# Stopping the misinformation: BNT162b2 COVID-19 vaccine has no negative effect on women’s fertility

**DOI:** 10.1101/2021.05.30.21258079

**Authors:** Myriam Safrai, Amihai Rottenstreich, Shmuel Herzberg, Tal Imbar, Benjamin Reubinoff, Assaf Ben-Meir

## Abstract

**Objective:** To investigate the possible impact of Pfizer-BioNTech’s mRNA BNT162b2 COVID-19 vaccine on women’s fertility.

**Methods:** A retrospective single-center study examining women’s IVF treatment parameters and pregnancies before and after their vaccination between February and May 2021. Each woman served as a self-control before and after vaccination. Additionally, in order to neutralize the effect of the sperm on fertilization, only Intracytoplasmic Sperm Injection (ICSI) patients who were currently being treated with an ICSI cycle and had an earlier ICSI cycle available were included in the study. The study outcomes compared between the PRE and POST vaccination groups and consisted of: the IVF cycle outcomes, including the number of oocytes retrieved; the number of matured oocytes; the fertilization rate; and the number and quality of embryos at day 3. Clinical pregnancy was based on the first hCG value reported if the data were available for both cycles.

**Results:** A final total of 47 women were eligible for inclusion with a mean interval of 362 ±368 days between the two ovum pick ups. The characteristics of their ICSI cycles before and after the vaccination were similar for all the parameters. Additionally, the number and percentage of clinical pregnancies did not significantly differ between the PRE and POST vaccination groups (n=15).

**Conclusion:** This study is the first to evaluate the impact of the BNT162b2 vaccine on women’s fertility. From our findings, the vaccine appears to have no impact on women’s fertility. This study is the first step in abolishing the misinformation derived from unreliable sources and reassuring patients in order to improve compliance and promote COVID-19 eradication.

## Introduction

Severe acute respiratory syndrome coronavirus 2 (SARS-CoV-2), responsible for causing Coronavirus Disease 19 (COVID-19), has affected over 160 million of people worldwide since it was declared a pandemic in March 2020 by the World Health Organization (WHO)(1), and has caused over 3 million deaths (2). The resulting urgent need for effective tools to combat COVID-19 has led to the accelerated development and recent approval of the BNT162b2 mRNA vaccine launched by BioNTech and Pfizer (3). In a large trial, the two-dose regimen of BNT162b2 vaccine was assessed and found to confer 95% protection rate against SARS-CoV-2 among individuals aged 16 and older (4,5). Based on this information, a mass vaccination campaign using the BNT162b2 vaccine began in Israel (6) with the recommendation to vaccinate the entire population aged 16 and above (5). The safety profile of the aforementioned vaccine was previously confirmed assessed using self-reporting of local and systemic adverse events, the use of antipyretic or pain medication, and unsolicited serious adverse events (3,7). However, the effect of the vaccine on fertility has not yet been investigated.

Reproductive-aged women are considered a special population and often are not included in clinical trials (17). Indeed, pregnant women and women trying to conceive were excluded from the pivotal clinical trials evaluating the mRNA-based vaccines (3,8,9). This has resulted in many unanswered questions about the safety of the BNT162b2 vaccine on fertility. Since the launch of the vaccine, vaccination hesitancy has been a serious problem for COVID eradication (10). The social media panic has significantly increased the fear and hesitancy to receive the COVID-19 vaccine (11,12) in large parts of the population (10,13). The impact of the vaccine on fertility has also been the subject of many rumors and misinformation.

The adult female population has another unique and challenging aspect. Pregnant women are at a higher risk of complications and increased risk of perinatal complications, if they become infected with COVID-19 (9,14–20). Therefore the American Society of Reproductive Medicine Task Force does not recommend withholding the vaccine from patients who are planning to conceive (21). Due to the lack of information and the extreme clinical relevance, we aimed to investigate the possible impact of the BNT162b2 COVID-19 vaccine on women’s fertility.

## Methods

This study was carried out at the Hadassah Hebrew-University Medical Center, a university hospital with a large in vitro fertilization (IVF) unit in Israel. Data were collected from all patients treated in the IVF unit between February and May 2021 after the vaccination of the general population began. Medical records of patients who had received two doses of BNT162b2 vaccine were retrospectively reviewed (PRE-vaccine) using the hospital’s electronic database and were compared to prospectively collected data of those patients (POST-vaccine).

In order to minimize bias, each woman served as a self-control before and after vaccination. Additionally, in order to neutralize the effect of the sperm on fertilization, only Intracytoplasmic Sperm Injection (ICSI) patients who were currently being treated with an ICSI cycle and had an earlier ICSI cycle available were included in the study.

Data obtained included: patient demographics (age and body mass index (BMI)); indication for IVF treatment (i.e. female/male factor, unknown infertility, and need for pre-implantation genetic diagnosis (PGD) or fertility preservation); follicle stimulating hormone (FSH) value; data regarding the IVF cycle (length of gonadotropin (GT) stimulation and total GT dose, estrogen level on the day before ovum pick up (OPU), the number of oocytes retrieved, the number of mature oocytes, the number of fertilized oocytes, the number and quality of embryos at day 3); and the time since the first dose of the vaccine. The second vaccine dose was given as recommended, 21 days after the first dose (5,7). The embryos’ quality at day 3 were determined by cell number, symmetry and fragmentation according to the Society for Assisted Reproductive Technology (SART) grading guidelines and graded as good, fair or poor (22). Clinical pregnancy based on the first hCG value was reported if the data were available for both cycles. The pregnancy rate was calculated from the total of cycle with data for both cycles (n=15).

The primary outcomes compared between the PRE and POST vaccination groups and consisted of: the IVF cycle outcomes, including the number of oocytes retrieved; the number of matured oocytes; the fertilization rate; and the number and quality of embryos at day 3.

### Statistical analysis

IVF treatment parameters are presented as mean +/-standard deviation (SD) or percentage. Comparisons between pre- and post-vaccine values were conducted with paired t-tests. A Pvalue of 0.05 or less was considered significant. The significance of pregnancy rate before and after vaccination was assessed by McNemer’s test. All statistical analyses were carried out using Excel 2013. A sample size of 32 women (in the entire cohort) was required to detect a significant difference of 30% in the number of oocytes retrieved (probability of Type 1 error of 0.05 and 80% power).

### Ethical approval

Approval was obtained from the institutional review board (IRB) of the Hadassah-Hebrew University Medical Center before data extraction was performed. The requirement for written informed consent was waived by the IRB.

## Results

During the study period, 297 women were treated in the IVF unit. Most of them (n= 56%) had completed the two doses of the BNT162b2 vaccine. Thirteen percent (n=39) of the women had prior history of SARS-CoV-2 infection while 87% (n=258) were eligible for vaccination. In this subgroup of women who were eligible for vaccination, 64% had completed the two vaccine doses, 2% had received only one dose and 34% chose not to be vaccinated for COVID-19 (Figure 1).

**Figure 1.**
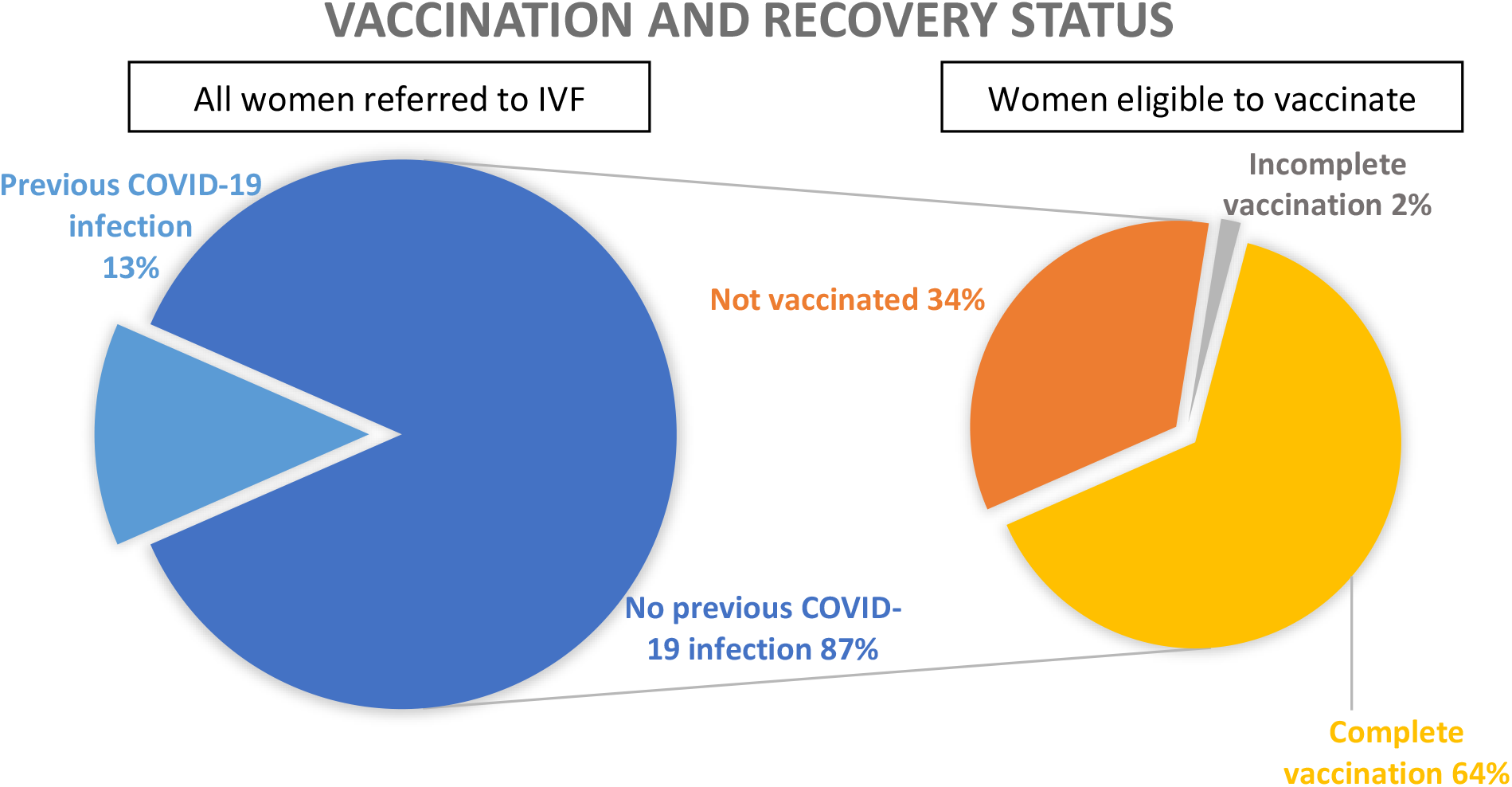
Vaccination and recovery status of women treated in the In Vitro Fertilization clinic. The figure shows (left side) that 13% of the women referred to the IVF clinic during the study period were previously infected with COVID-19 and were ineligible for vaccination. In the remaining 87% of women (right side): 64% were fully vaccinated, 2% had completed a single dose of the vaccine, and 34% were unvaccinated.

A flow chart describing patient inclusion in this study is shown in Figure 2. The final cohort included atotal of 47 women and their demographic data and indications for undergoing IVF are shown in Table 1. These women had a mean interval of 362 ±368 days between the two OPU. The characteristics of their IVF cycles before and after the vaccination were similar for all the parameters (Table 2).

**Figure 2.**
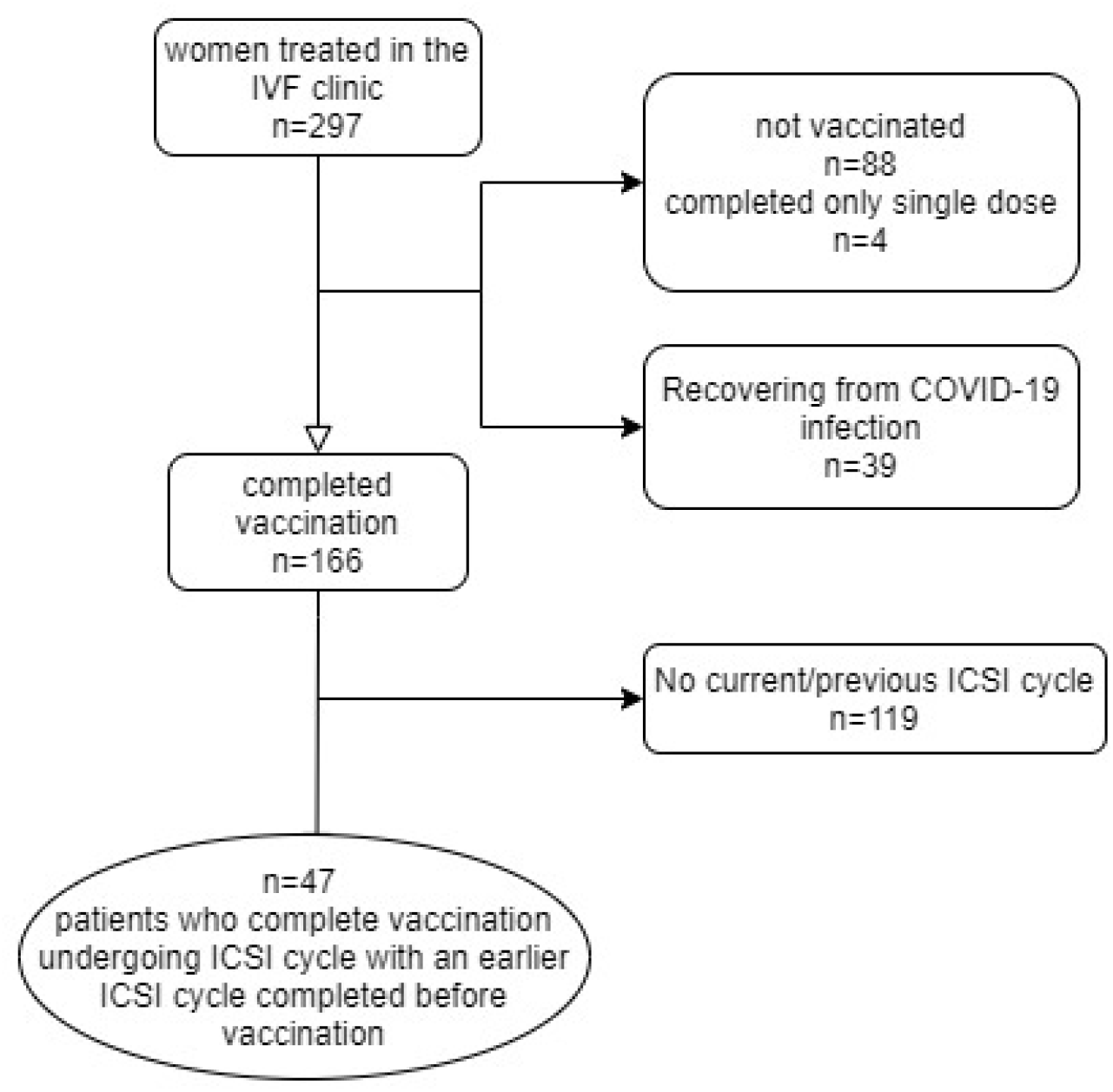
Study design IVF – In Vitro Fertilization, ICSI - Intracytoplasmic Sperm Injection.

**Table 1.** Demographics of patient sample and indication for IVF

|  |  |
| --- | --- |
| Age (years) | 37.36 ± 7.5 |
| Body Mass Index (BMI) | 27.5±6.2 |
| Time from the first vaccine dose (days) | 57.3 ± 24.7 |
| FSH (IU/L) | 9.27 ± 4.9 |
| Interval between both OPU (days) | 362.7 ± 386.4 |
| **Indication for IVF (%)** |  |
| Female | 49 |
| Male | 19 |
| Unexplained | 21 |
| PGD | 11 |

All continuous variables are expressed as mean ± standard deviation (SD)

**Table 2.**
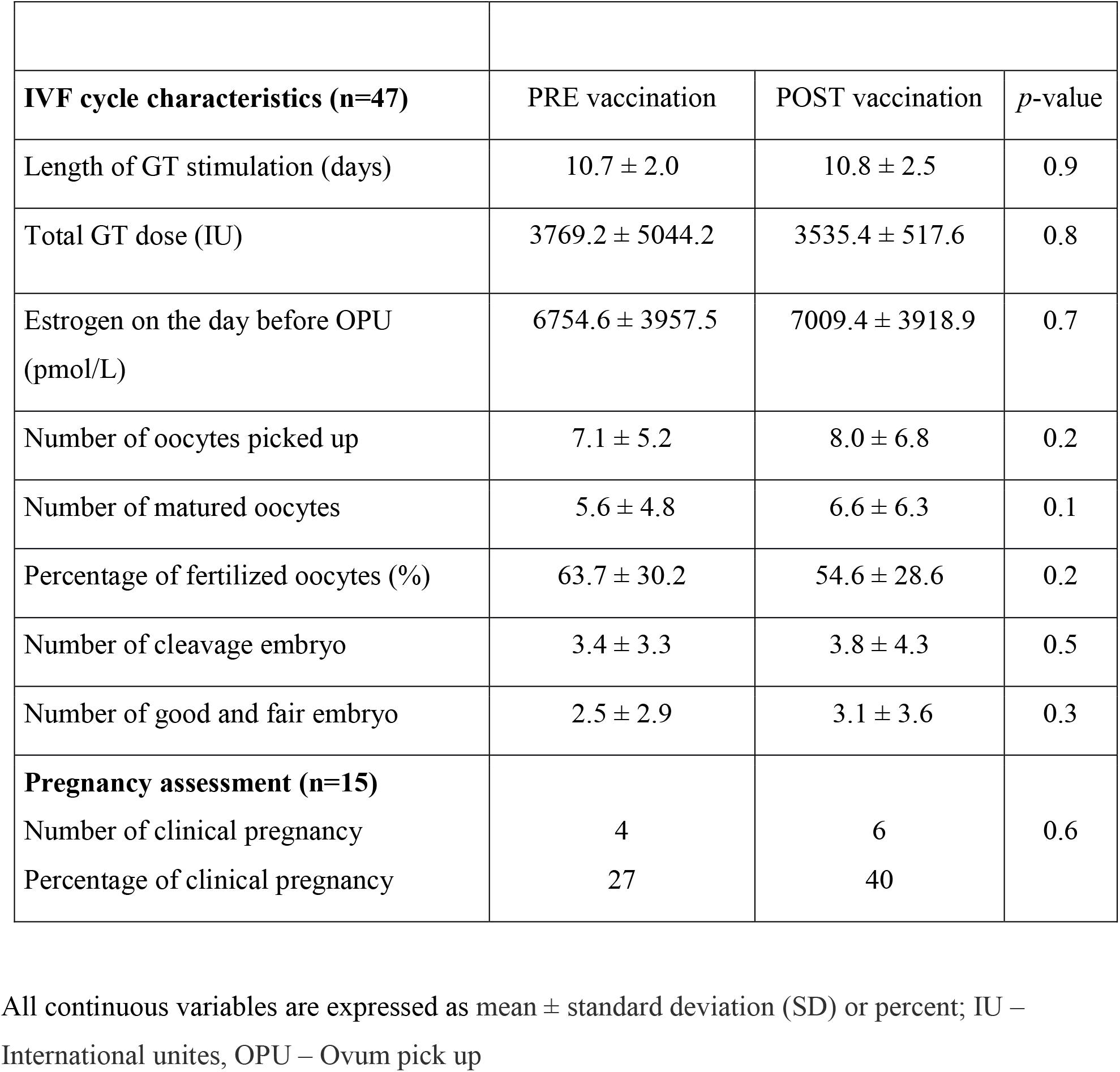
Patients’ IVF cycle parameters and outcomes before and after BNT162b2 vaccination

| **IVF cycle characteristics (n=47)** | PRE vaccination | POST vaccination | *p*-value |
| --- | --- | --- | --- |
| Length of GT stimulation (days) | 10.7 ± 2.0 | 10.8 ± 2.5 | 0.9 |
| Total GT dose (IU) | 3769.2 ± 5044.2 | 3535.4 ± 517.6 | 0.8 |
| Estrogen on the day before OPU (pmol/L) | 6754.6 ± 3957.5 | 7009.4 ± 3918.9 | 0.7 |
| Number of oocytes picked up | 7.1 ± 5.2 | 8.0 ± 6.8 | 0.2 |
| Number of matured oocytes | 5.6 ± 4.8 | 6.6 ± 6.3 | 0.1 |
| Percentage of fertilized oocytes (%) | 63.7 ± 30.2 | 54.6 ± 28.6 | 0.2 |
| Number of cleavage embryo | 3.4 ± 3.3 | 3.8 ± 4.3 | 0.5 |
| Number of good and fair embryo | 2.5 ± 2.9 | 3.1 ± 3.6 | 0.3 |
| **Pregnancy assessment (n=15)** |  |  |  |
| Number of clinical pregnancy | 4 | 6 | 0.6 |
| Percentage of clinical pregnancy | 27 | 40 |  |

All continuous variables are expressed as mean ± standard deviation (SD) or percent; IU – International unites, OPU – Ovum pick up

Various outcomes of the IVF cycle were found to be not significantly different before and after vaccination, such as the number of oocytes retrieved (7.1 ±5.2 vs. 8.0 ±6.8, p=0.2), the number of mature oocytes (5.6 ±4.8 vs. 6.6 ±6.3, p=0.1) and the fertilization rate (63.7±30.2 vs. 54.6 ±28.6, p=0.2). In addition, embryos parameters were assessed and no significant differences were found between the number of cleavage embryos (3.4 ±3.3 vs. 3.8 ±4.3, p=0.5) and the number of good and fair embryos (2.5 ±2.9 vs. 3.1 ±3.6, p=0.3) before and after vaccination.

An assessment of the pregnancy rate was carried out on 15 women from the sample. The number and percentage of clinical pregnancies did not significantly differ between PRE and POST vaccination groups (Table 2). Three women were pregnant in both cycle, one woman was pregnant only in her earlier cycle compared to two women pregnant after their vaccination. Nine women did not conceive in any of their cycles, resulting in a non-significant difference of p=0.56.

## Discussion

This study is the first to evaluate the impact of the BNT162b2 vaccine on women’s fertility. From our findings, the vaccine appears to have no impact on women’s fertility. Specifically, no differences were found between the ICSI cycles that each patient underwent before and after vaccination. All the ICSI outcomes including: the number of oocytes retrieved, the number of matured oocytes and the percentage of fertilized oocytes were similar in the PRE and POST vaccination groups. Moreover, we continued the follow up for a few more days and assessed the quantity and quality of cleavage embryos and found no changes. Lastly, a subgroup from our sample showed that the pregnancy rate was also similar in the PRE and POST vaccination groups. These findings therefore are the first step in showing that the BNT162b2 vaccine has no effect on IVF treatment parameters nor on the pregnancy rate. Our study results thus support the statement from the American College of Obstetricians and Gynecologists, the American Society for Reproductive Medicine, and the Society for Maternal-Fetal Medicine that “no loss of fertility has been reported among trial participants or among the millions who have received the vaccines since their authorization, and no signs of infertility appeared in animal studies. Loss of fertility is scientifically unlikely” (23).

The impetus for this study came about due to the current misinformation and rumors widely disseminated on social media of the effects of the BNT162b2 vaccine on women’s fertility. This seems to have an effect on vaccination compliance. Of the 297 women treated in the IVF unit during the study period, only 56% had undergone full vaccination i.e. two doses of the BNT162b2 vaccine. This is much lower than Israel’s national average vaccination rate of 71-80% in the general population aged 20-49 years (24). Such fears may be based on various non-scientifically sound claims (9), which reinforce the normal uneasiness felt in response to a new vaccine. One of the baseless arguments for the BNT162b2 vaccine negatively impacting women’s fertility was that the vaccine contains a spike protein called syncyntin-1, which is vital also for the formation of the placenta. Antibodies produced against this protein may attack the placenta too, leading to abortions. However, these claims have since been revoked as the vaccine contains neither syncyntin-1 nor the mRNA sequence for syncyntin-1(21).

Another possible claim for the BNT162b2 vaccine causing female infertility is that the functional receptor for SARS-CoV-2 is ACE2 (Angiotensin converting enzyme-2), which modulates the cleavage of angiotensin II in the renin-angiotensin system. COVID-19 invades host cells and downregulates ACE2 expression causing an increased proinflammatory response by angiotensin II. Since ACE2 and angiotensin II regulate basic reproductive functions such as folliculogenesis, steroidogenesis, oocyte maturation and ovulation, there was the concern that the vaccine which mimics the virus could also reduce fertility by the same mechanism(25). However this has not been proven and moreover, the BNT162b2 vaccine does not have the ACE2 receptor to cause such infertility (26). Our results also refute this claim since we found there was no negative impact on folliculogenesis and embryogenesis, as the number of oocytes and their maturation was not impaired.

Our study has several limitations. The main one is the retrospective nature of the analysis of the PRE vaccine group. This carries an inherent selection bias and information bias due to the medical record coding. Another potential caveat is the relatively small sample size of the study population. Nevertheless, the sound methodology where each patient served as their own self-control strengthens the results and allows us to provide reliable answers to the questions raised regarding the effects of BNT162b2 vaccination on fertility. Finally, we included data on the pregnancy rate in a subgroup of the study sample (n=15). This is a small group but we included this data as it strengthens our findings showing no differences in IVF treatment parameters before and after vaccination and moreover it directly analyzes women’s fertility. There is an inherent bias in considering the pregnancy rate; couples whose previous ICSI cycle ended in pregnancy are less likely to return for another cycle. The mean interval between both OPU was 362 days. The impact of such a period of time on fertility differs depending on the age of the patient; namely, it is more significant in older women. However, no such fertility differences were seen in our study. If any such differences had in fact shown, we would expect reduced fertility of the POST group. Clearly, a larger study is warranted to validate these initial findings demonstrating that the BNT162b2 vaccination does not impact women’s fertility.

In conclusion, our study is the first to assess the impact of the BNT162b2 COVID-19 vaccine on women’s fertility, and provides encouraging data showing that this vaccine likely does not impair women’s fertility. This study is the first step in abolishing the misinformation derived from unreliable sources and reassuring patients in order to improve compliance and promote COVID-19 eradication.

## Data Availability

The data that support the findings of this study are available from the corresponding author, MS, upon reasonable request.

## Acknowledgement

The authors wish to thank Dr. Ruth Moont for her contribution to the reviewing and editing of the manuscript.

## Notes

Conflict of interest: The authors have declared that no competing interests exist

### Competing Interest Statement

The authors have declared no competing interest.

### Funding Statement

The authors received no specific funding for this work

### Author Declarations

The study was approved by the institutional review board, Hadassah Medical Center IRB (Permit 0054-21-HMO).

